# Comparative Evaluation of Wearable Sensor Form Factors for Physiological Monitoring in Youth with Autism Spectrum Disorder

**DOI:** 10.64898/2026.05.06.26352564

**Authors:** Caden Stewart, Abigail Albertazzi, Jacqueline Tasarz, Kristen Kim, Veronica Gandara, Corrine Blucher, Camila C. Reyes-Martinez, Benjamin Smarr, Aaron D. Besterman

## Abstract

Sudden behavioral outbursts in youth with autism spectrum disorder (ASD) are difficult to predict and create substantial caregiving burdens. Wearable physiological monitoring might enable prediction, but sustained use may be limited by tolerability. We evaluated adherence and data completeness in 40 youth with ASD over a two-week period across four device types (wristband, headband, adhesive chest patch, and finger ring) alongside caregiver-reported useability and comfort. Data completeness varied markedly by device, with the patch achieving the highest completeness (∼80%), followed by the wristband (∼60%), headband (∼50%), and ring (∼20%). In multivariate analyses, adherence was driven by the device form factor rather than participant-level clinical characteristics. Devices rated as more comfortable did not yield higher completeness, revealing a divergence between reported preference and actual use. These findings suggest that device choice is a key consideration for studies in ASD youths, highlighting the need for research into model stability across sensor types in neurodivergent populations.

## Introduction

Behavioral outbursts (e.g., aggression, self-injury, or elopement) are among the most distressing and functionally impairing challenges for youth with autism spectrum disorder (ASD) and their families^1,2^. Behavioral outburts often arise abruptly, disrupt learning and social functioning, strain caregiver relationships, and lead to emergency service utilization or hospitalization^3^. Caregivers consistently identify behavioral outbursts as one of the most overwhelming aspects of caring for autistic youth^4,5^, yet clinical tools for anticipating or preventing these events remain extremely limited.

A growing body of work suggests that behavioral outbursts are not purely sudden events but rather the endpoint of an escalating physiological process^6–10^. Studies using wearable biosensors have shown that autonomic signals such as heart rate and electrodermal activity often change in the minutes preceding aggressive or dysregulated behavior in autistic youth^6–9^. These findings suggest that continuous physiological monitoring could provide an objective window into escalating arousal that is difficult to detect through observation alone. More broadly, research in other episodic psychiatric conditions, including panic^11^ and bipolar disorder^12^, demonstrates that wearable physiological signals such as heart rate, heart rate variability, sleep disruption, and circadian instability can signal increasing vulnerability hours or days before symptom onset.

Despite this potential, translation of wearable monitoring into real-world autism care has been slow. Two critical and related hurdles are device tolerability and adherence. Many autistic children experience sensory sensitivities that can make continuous wearable use difficult^13^. As a result, data loss and inconsistent wear remain major limitations in wearable research involving autistic individuals^14^. Importantly, most prior studies have evaluated only a single wearable platform, making it difficult to determine whether feasibility limitations reflect inherent challenges of wearable monitoring in ASD or simply the characteristics of the specific device used^10^. This has hindered the development of reliable digital biomarkers and limited the reproducibility of findings across studies. Consequently, a critical unanswered question is which wearable form factors can reliably support continuous physiological monitoring in autistic youth for both predictive model building and, ultimately, intervention. Establishing this foundation is essential for advancing the next generation of digital health research in ASD. Without identifying devices that are both tolerable and capable of generating high-quality physiological data over extended periods, efforts to develop predictive algorithms or real-world monitoring systems will remain constrained by incomplete or biased datasets. Systematic evaluation of wearable platforms is therefore a necessary step toward establishing best practices for digital phenotyping in neurodevelopmental populations.

The present study addresses this gap through a pilot trial comparing four wearable form factors— a finger ring, wristband, chest patch, and headband—in youth with ASD. Over a two-week monitoring period, we evaluated device tolerability, adherence, and data completeness while integrating caregiver feedback regarding usability and burden. This comparative design allows us to identify which sensing platforms are most compatible with the sensory, behavioral, and practical realities of daily life for youth with ASD. By identifying wearable configurations that can support sustained, high-quality physiological data collection in this population, this work establishes a methodological foundation for future digital health studies in ASD. These findings can inform device selection, study design, and data collection strategies for subsequent research aimed at developing predictive models of behavioral dysregulation and just-in-time interventions^15^. More broadly, defining practical standards for wearable monitoring in youth with ASD represents a critical step toward translating physiological sensing technologies into scalable tools for early detection and prevention of behavioral crises.

## Results

### Enrollment

A total of 96 youth with ASD were assessed for eligibility through electronic health record screening, of whom 40 were enrolled and assigned to one of four wearable device groups (Fig. 1). Enrollment proceeded in a staged manner, with exclusions primarily due to inability to contact families, scheduling constraints, or lack of interest following prescreening. Of those enrolled, 32 participants completed the full monitoring protocol and were included in questionnaire-based analyses, while 28 provided sufficient device data for adherence analyses. Attrition occurred both prior to device initiation and during the monitoring period, most commonly due to device intolerance or loss to follow-up (Fig. 1).

**Figure 1.**
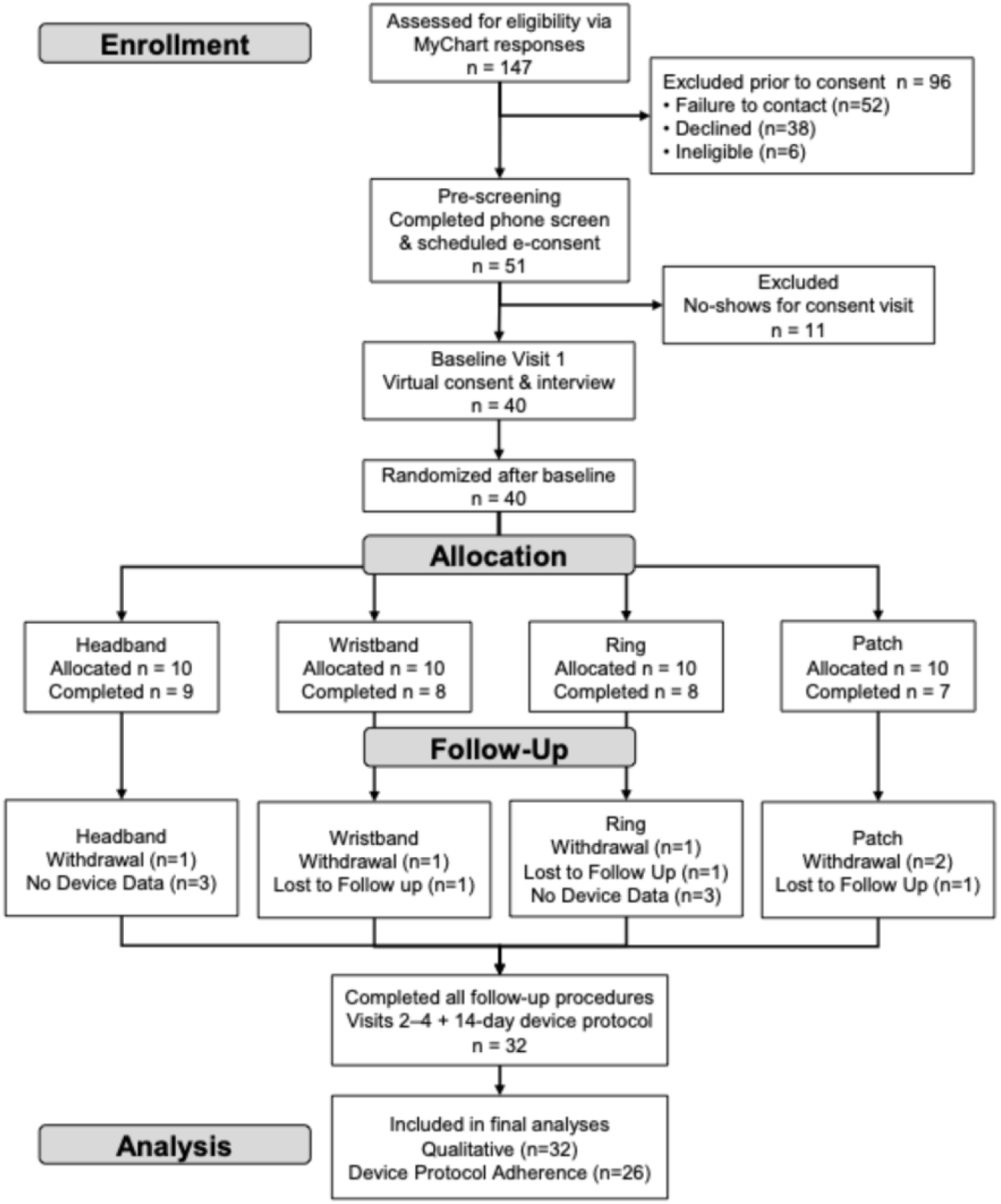
A participant flow diagram showing the progress through each stage of the present study, including enrollment, device group allocation, follow-up procedures, and final analyses.

### Adherence

Adherence to the wearable device protocol varied substantially across device form factors (Fig. 2). The adhesive chest patch (VivaLNK) demonstrated the highest adherence, with approximately 80% data completeness. The wrist-worn device (Fitbit) showed intermediate adherence (∼60% data completeness), whereas devices intended primarily for nighttime use (Muse headband and Wellue ring) exhibited lower data capture overall, even after accounting for shorter expected recording windows (∼50% and 20%, respectively). Among these, the Muse headband yielded greater data availability than the ring, with average nightly use of approximately 4 hours. Across all device groups, adherence exhibited considerable inter-individual variability.

**Figure 2.**
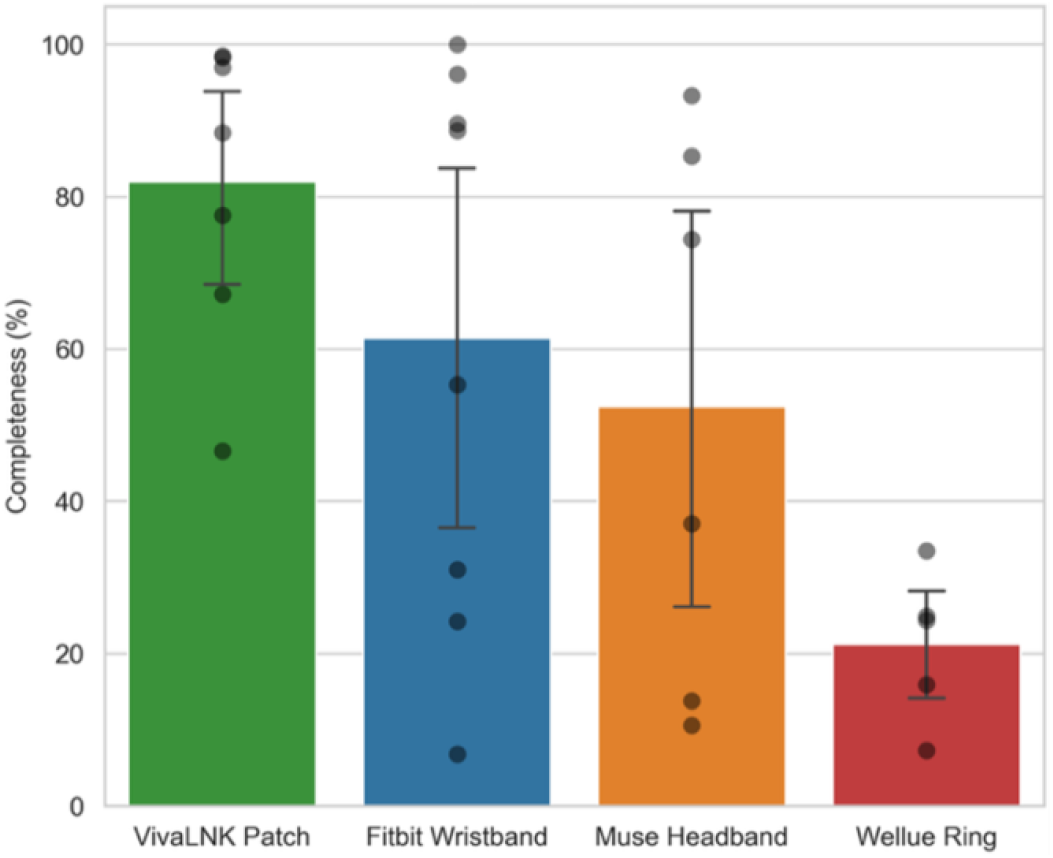
Data completeness across wearable device form factors.Mean completeness (%) over the two-week monitoring period by device, with 95% CI (error bars) and individual participants (points). Completeness reflects the proportion of expected recording time with valid data.

Temporal patterns of adherence display day-level completeness for daytime wear periods across participants (Fig. 3). These visualizations demonstrate consistently higher adherence for the adhesive patch compared to the wristband, as well as substantial inter-individual variability. Across both devices, adherence was highest at the beginning of the monitoring period and declined over time.

**Figure 3.**
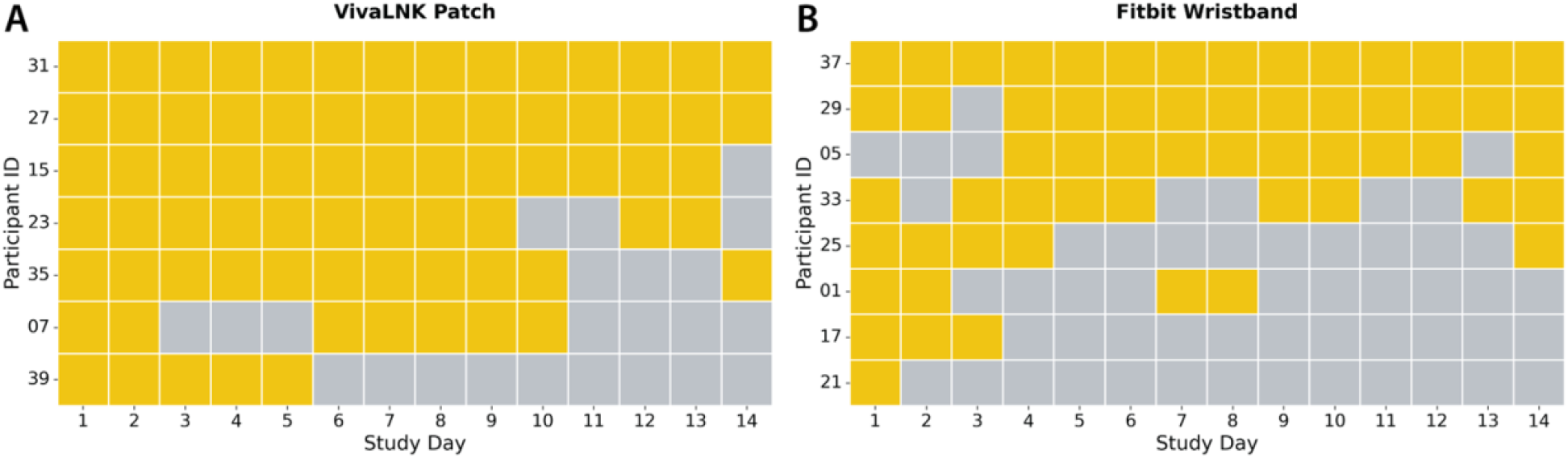
Daytime adherence across wearable device form factors. Complete daytime periods over the two-week monitoring period by device. Daytime was defined as 08:00–22:00; a period was considered complete if ≥10 hours of valid data were recorded.

Nighttime adherence patterns across all devices demonstrated consistently high adherence across participants for the patch, with more complete nighttime coverage than any other device, including those designed specifically for sleep monitoring (Fig. 4). In contrast to daytime patterns, adherence at night was less characterized by gradual decline and instead showed more intermittent, participant-specific variability. The patch also exhibited a higher floor of use, with all participants contributing multiple nights of data, whereas other devices included participants with minimal or no recorded nights.

**Figure 4.**
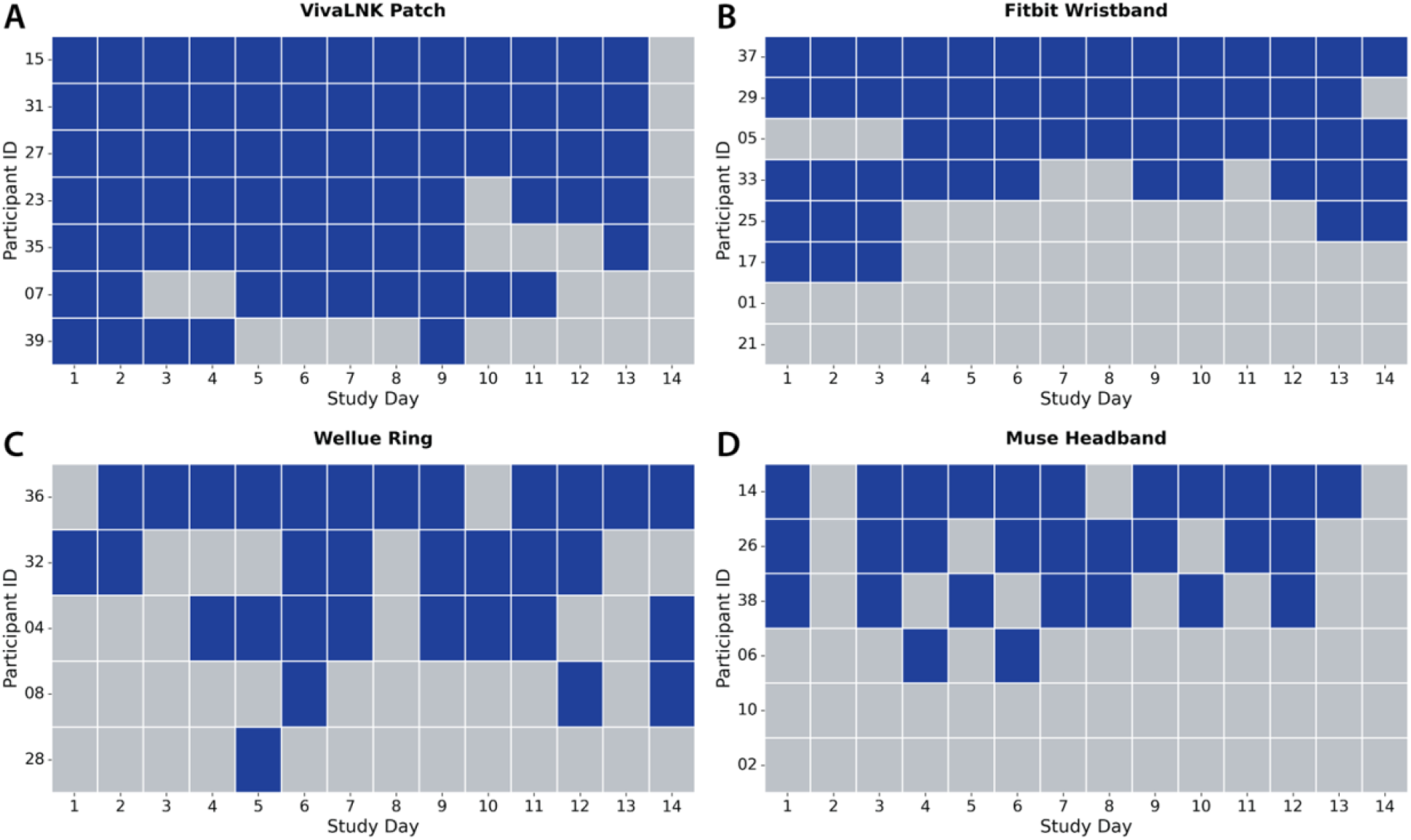
Nighttime adherence across wearable device form factors. Complete nighttime periods over the two-week monitoring period by device. Nighttime was defined as 22:00–08:00; a period was considered complete if ≥6 hours of valid data were recorded.

### Determinants of Wearable Protocol Adherence

Table 1. summarizes the results of a linear regression model evaluating predictors of overall data completeness across the two-week monitoring period. Regression coefficients (β) represent the estimated change in completeness associated with each predictor. The overall model demonstrated marginal significance (p = 0.049), suggesting limited explanatory value. Clinical variables and biological sex were not significant predictors of adherence and were associated with large standard errors, indicating substantial uncertainty in their estimates. In contrast, device type emerged as the primary determinant of adherence. Specifically, assignment to the ring device was associated with significantly lower data completeness relative to the wristband reference group. The intercept reflects the expected completeness for the reference group (participants assigned to the wristband with baseline covariates), while the significant negative coefficient for the ring indicates reduced adherence compared to this group. Overall, these findings suggest that variation in adherence was driven predominantly by device form factor rather than participant-level clinical characteristics.

**Table 1.** Linear regression model predicting data completeness.

| Predictor | B | SE | p |
| --- | --- | --- | --- |
| Intercept | 71.51 | 21.97 | .005** |
| **Clinical Indicators** |  |  |  |
| ADHD (Present) | -2.17 | 14.07 | .880 |
| OCD (Present) | 17.18 | 20.87 | .422 |
| Depression (Present) | -22.61 | 21.10 | .300 |
| Anxiety (Present) | 21.43 | 21.49 | .333 |
| **Demographics** |  |  |  |
| Sex (Male) | -16.13 | 15.66 | .318 |
| **Device Type** |  |  |  |
| Muse Headband | -15.62 | 17.77 | .392 |
| VivaLNK Patch | 20.78 | 16.97 | .239 |
| Wellue Ring | -45.09 | 18.38 | .026* |

Note. OLS regression. Coefficients (B) represent the estimated change in completeness relative to the reference group (wristband). SE, standard error. Model fit: $R^2 = 0.566$ , adjusted $R^2 = 0.348$ , $p = 0.049$ .
\* $p < 0.05$ , \*\* $p < 0.01$ .

### Comfort and Usability Scoring

Subjective usability and comfort were assessed using the System Usablity Scale (SUS)^16^ and Comfort Rating Scale (CRS)^17^. Usability scores were highest for the wrist-worn device (Fitbit), with lower scores observed for the ring, headband, and adhesive patch (Fig. 5). A similar pattern was observed for comfort, with the Fitbit demonstrating the highest mean CRS scores and the other devices showing lower and more variable ratings. Although the adhesive patch demonstrated superior adherence, it was associated with comparatively lower usability and comfort scores. Despite these differences in mean values, group-level comparisons were not statistically significant for either usability (SUS; p = 0.07) or comfort (CRS; p = 0.61).

**Figure 5.**
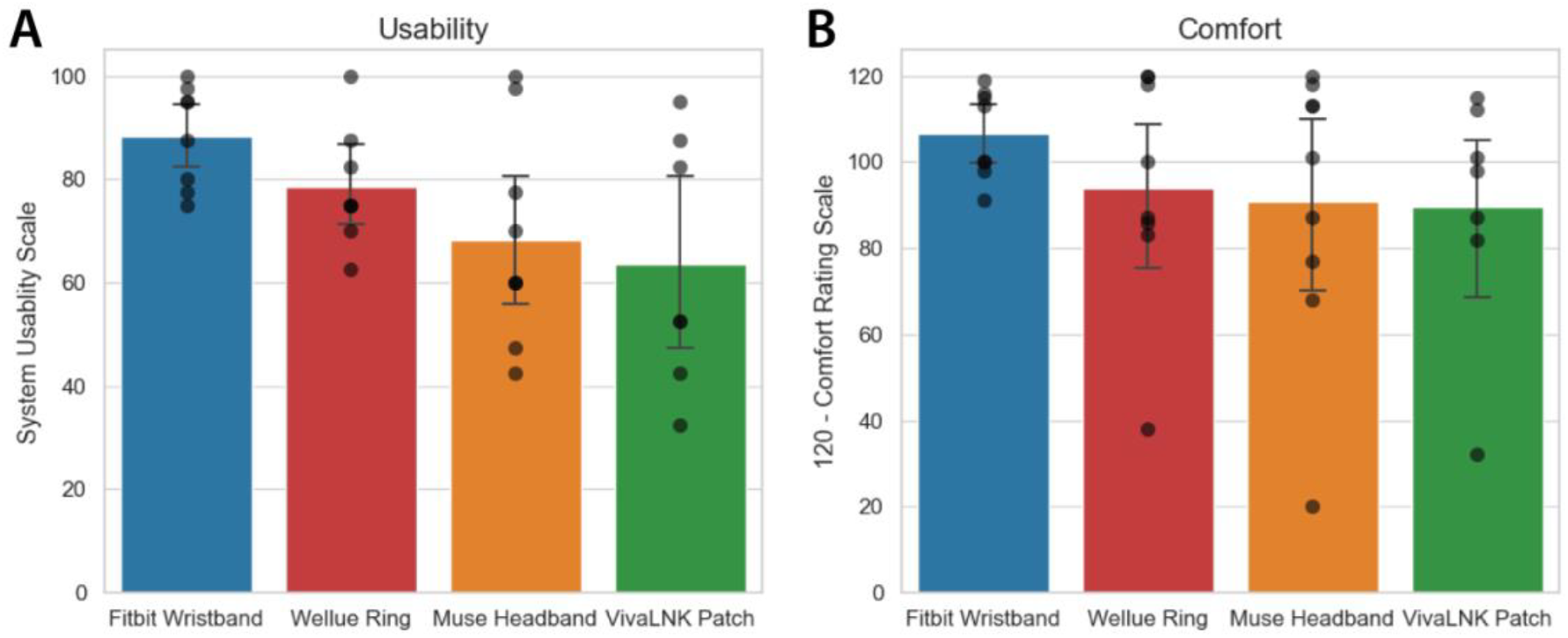
Usability and comfort scores across wearable device form factors. (A) Mean System Usability Scale (SUS) and (B) Comfort Rating Scale (CRS) scores by device over the two-week monitoring period. Error bars represent 95% confidence intervals, and individual data points are shown.

### Exploration of Dimensional Behavioral Symptoms

Given the clinical heterogeneity of ASD, we explored whether variation in device adherence was associated with caregiver-reported dimensional behavioral symptoms. Linear regression analyses were performed to assess the relationship between CBCL total scores (broad emotional and behavioral problems) and SRS-2 scores (autistic trait burden) and overall data completeness across the monitoring period (Fig. 6). There was no meaningful association between either measure and adherence (CBCL: R^2^ = 0.01; SRS-2: R^2^ ≈ 0.00), suggesting that variation in behavioral symptom burden, including autistic traits and broader emotional/behavioral problems, was not a major determinant of device adherence in this cohort.

**Figure 6.**
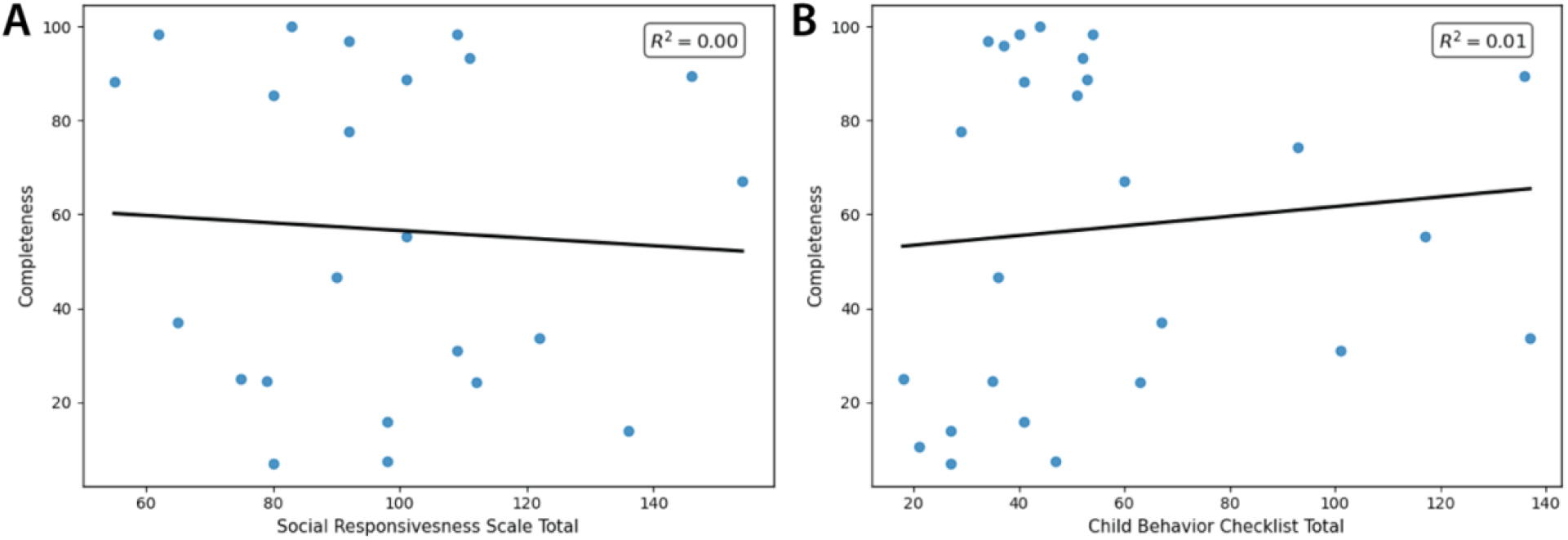
Relationship between dimensional behavioral symptoms and device adherence. Linear regression of data completeness (%) against Social Responsiveness Scale–2 (SRS-2) scores (A) and Child Behavior Checklist (CBCL) total scores (B). Lines represent ordinary least squares fits; R^2^ values are shown for each model.

### Qualitative Responses

Semi-structured caregiver interviews provided additional context for adherence patterns. Reported benefits centered on access to physiological data and its role in supporting health awareness and parent–child communication, particularly for devices providing sleep and activity metrics. Common challenges included device management (e.g., charging, syncing), physical discomfort, and disruption to routines, with these issues varying by form factor. Notably, devices requiring frequent charging or nighttime-only wear were associated with more reported challenges, whereas the adhesive patch was primarily limited by practical considerations such as adhesive and moisture exposure.

## Discussion

Our findings highlight the need to prioritize user-centered design and deployment strategies in neurodevelopmental populations. Wearable technologies are often evaluated primarily on their sensing capabilities, yet our results suggest that form factor and usability may be equally, if not more, important for successful real-world implementation. These results extend a growing body of work demonstrating the promise of wearable biosensing for predicting clinically meaningful behaviors in ASD. Prior studies have shown that physiological and motion signals can anticipate aggressive or dysregulated behaviors with moderate-to-high accuracy. For example, Goodwin et al. demonstrated that wrist-worn biosensors could predict aggression up to one minute before onset in naturalistic inpatient settings^4^, and subsequent multi-site work replicated and extended these findings, achieving predictive performance with AUROC values around 0.80^5^. Similarly, laboratory-based studies have shown that increases in heart rate can precede challenging behaviors in young children with ASD^6^, supporting the biological plausibility of anticipatory physiological signals. Feasibility studies have further demonstrated that certain wearable devices can achieve acceptable comfort and data robustness under controlled conditions^7^.

However, these studies have largely focused on prediction performance under conditions optimized for data collection, including structured inpatient environments, intensive observation protocols, or laboratory paradigms. In contrast, our study shifts attention to a more fundamental and underexamined question: whether such data can be reliably obtained in real-world, naturalistic settings. While prior work has established that wearable signals can be predictive when available, we show that the availability of usable data itself is highly dependent on device form factor. In this context, variability in wearable datasets is not only an analytic challenge but also a practical constraint: device selection directly influences whether sufficient data are captured to enable downstream modeling at all.

A key implication is that data completeness represents a critical translational bottleneck for wearable-based digital health systems. Continuous monitoring and just-in-time adaptive interventions rely on sustained, high-quality data streams. While prior studies have demonstrated the feasibility of predicting behavior from physiological signals, our findings suggest that the real-world reliability of such systems will depend as much on sustained data capture as on predictive model performance. In this sense, data completeness functions as a prerequisite for valid inference and clinical utility rather than a secondary technical consideration.

A particularly important finding is the divergence between data completeness and user experience across device types. While the patch-based device yielded the highest data completeness, caregiver-reported usability and comfort were highest for the wristband and lowest for the patch. Qualitative feedback from caregivers further supports this pattern, highlighting differences in tolerability, ease of use, and day-to-day feasibility across form factors. Together, these findings suggest a fundamental tradeoff: devices that optimize continuous physiological data capture may not be those that are most acceptable for sustained real-world use.

This tradeoff has important implications for the design and deployment of wearable-based systems. One potential approach is to decouple data collection from implementation, using higher-fidelity devices (e.g., patches) for model development and training, while deploying more acceptable devices (e.g., wristbands) for real-world monitoring and intervention. However, this strategy introduces a critical requirement: predictive models must generalize across device types. Differences in sensor placement, signal quality, and user behavior may degrade performance generalizability if not explicitly addressed. Ensuring cross-device robustness will therefore be essential for translating wearable biosensing from controlled studies to scalable clinical applications.

From a methodological perspective, these findings underscore the importance of explicitly measuring and reporting data completeness in wearable studies. Missingness is often treated as a nuisance or addressed through post hoc imputation; however, in real-world deployments, incomplete data may reflect interactions between device design and user experience. Incorporating completeness metrics as primary outcomes, and systematically evaluating their determinants, may be essential for advancing wearable-based research from feasibility to scalable implementation. Designing flexible, tolerable, and context-appropriate devices will likely be critical to ensuring that wearable systems can generate reliable data across diverse users.

Several limitations should be considered. First, the sample size was modest, which limits statistical power and the precision of estimates for participant-level predictors. Second, the study evaluated a specific set of device form factors in a defined deployment context, and findings may not generalize to other sensor platforms or study designs. Third, dimensional behavioral measures (SRS-2 and CBCL) were examined in exploratory analyses and may not fully capture other relevant factors influencing adherence, such as sensory sensitivities or user preferences. Future studies incorporating larger samples, richer phenotyping, and direct measures of user experience will be important for clarifying these relationships.

In conclusion, our findings suggest that the success of wearable biosensing in ASD depends not only on advances in signal processing and predictive modeling, but also on the ability to reliably capture data in real-world settings. Device form factor emerges as a key determinant of adherence, emphasizing that human-device interaction is central to the translation of wearable technologies into clinical practice. Future work should focus on integrating continuous physiological monitoring with prospective, real-world outcome prediction across devices and settings, enabling the development of scalable, personalized, and clinically actionable systems.

## Methods

### Participants

Forty adolescents with confirmed ASD and forty caregivers (one per adolescent) were serially enrolled to participate in the single site, non-randomized comparative feasibility study evaluating multiple wearable sensor form factors for physiological monitoring in autistic youth. The study protocol was reviewed and approved by the University of California, San Diego Institutional Review Board (UCSD IRB Protocol #804905). A partial waiver of HIPAA authorization was granted to allow prescreening of potentially eligible participants through the health system’s electronic medical record.

Potential participants were identified through electronic health record queries for adolescents aged 13–17 years with a documented diagnosis of ASD. Recruitment messages describing the study were sent to eligible families, and those who expressed interest were contacted by the study team for screening and eligibility assessment. Inclusion criteria required a confirmed historical ASD diagnosis established using the Autism Diagnostic Interview–Revised and the Autism Diagnostic Observation Schedule, Second Edition^21,22^. Exclusion criteria were minimal and limited to adolescents experiencing acute psychiatric instability, including active suicidal or homicidal ideation, that would make study participation unsafe.

Eligible youth and their caregivers attended an initial study visit during which study procedures, requirements, and potential risks were reviewed by trained research staff. Caregivers provided electronic informed consent, and youth provided electronic assent prior to participation. The enrolled adolescent participants ranged in age from 13 to 17 years (M = 15.06, SD = 1.30) and were predominantly male (70%), White (67.5%), and non-Hispanic (75%). Participants were sequentially allocated to one of four wearable device groups in a rotating order of enrollment to maintain balanced representation across device form factors. Eight participants withdrew after completing the initial consent visit and prior to receiving an assigned device or completing study procedures. For participants who continued in the study, data collection occurred across four research visits alternating between in-person and virtual formats, with the two-week wearable monitoring period occurring between Visit 2 and Visit 3 (Fig.1). The study was registered on ClinicalTrials.gov (NCT07504224).

### Data Collection

This study was conducted as a single-site comparative feasibility evaluation of wearable sensor form factors in youth with ASD. Study procedures occurred across four research visits that alternated between virtual and in-person formats. The two-week wearable monitoring period occurred between Visit 2 and Visit 3. Electronic caregiver consent and adolescent assent were obtained during Visit 1. Following enrollment, participants were sequentially assigned to one of four wearable device groups in a rotating order of enrollment to ensure balanced representation across device form factors.

Visit 2 was conducted in person and included baseline assessments and device onboarding. Youth completed the NIH Toolbox Cognition and Emotion assessment batteries^23,24^, while caregivers completed a brief medical history questionnaire as well as the Child Behavior Checklist (CBCL)^25^ and Social Responsiveness Scale–2 (SRS-2)^26^. Participants were then introduced to their assigned wearable device. To facilitate tolerability and reduce novelty-related anxiety, participants viewed a brief video social story describing the device, including its appearance, how it would be worn, and the types of physiological data collected during the two-week monitoring period. After viewing the video, study staff fitted the device to the youth and confirmed proper placement. The corresponding mobile application was installed on the participant’s or caregiver’s smartphone, and successful device synchronization was verified before participants left the visit. Participants received $20 compensation at the conclusion of Visit 2.

Participants were instructed to wear the device for two weeks during typical daily activities, removing it only when necessary, according to device-specific instructions. Two brief phone calls with caregivers occurred during the monitoring period—one after the first day of use and one after one week—to confirm device tolerability and successful data synchronization. The primary outcome of interest during the monitoring phase was completeness of physiological data capture, which served as an objective proxy for device adherence and tolerability. Visit 3 was conducted virtually following completion of the monitoring period. During this visit, adolescents and caregivers provided quantitative and qualitative feedback regarding device usability and comfort. Standardized usability measures included the System Usability Scale (SUS)^16^ and the Comfort Rating Scale (CRS)^17^. Participants also completed a brief semi-structured interview regarding their experience with the wearable device. Interview questions explored overall experience, perceived benefits, challenges of device use, perceived effects on mental well-being, and any additional feedback related to wearing the device. Responses were recorded by study staff. Visit 4 concluded participation. Adolescents returned their assigned wearable device during an in-person visit and received the final $20 compensation.

### Device Details

Four commercially available wearable devices representing distinct form factors were evaluated in this study: a wrist-worn device, headband, adhesive chest patch, and finger ring. All devices are approved for commercial use and were considered minimal risk. Potential risks primarily involved sensory discomfort associated with wearing the device, which may be particularly relevant for adolescents with autism spectrum disorder (ASD). These risks were explained during the informed consent process and were of particular interest in the study, as device tolerability represents an important factor influencing the feasibility of wearable physiological monitoring in this population. The devices evaluated included the Fitbit Inspire 2 wrist-worn activity tracker (FitBit, San Francisco, CA USA), the Muse S headband (InteraXon Inc, Toronto, ON, Canada), the VivaLNK VV350 Multi-Vital ECG Patch adhesive chest sensor (Vivalink, Campbell, CA, USA), and the Wellue O_2_ Ring finger-worn pulse oximeter (Wellue Health, Diamond Bar, CA, USA). Each device connected via Bluetooth to a corresponding mobile application installed on the participant’s smartphone, enabling continuous recording and synchronization of physiological data while the device was worn. Device characteristics and sensing capabilities are summarized in Table 2.

**Table 2.**
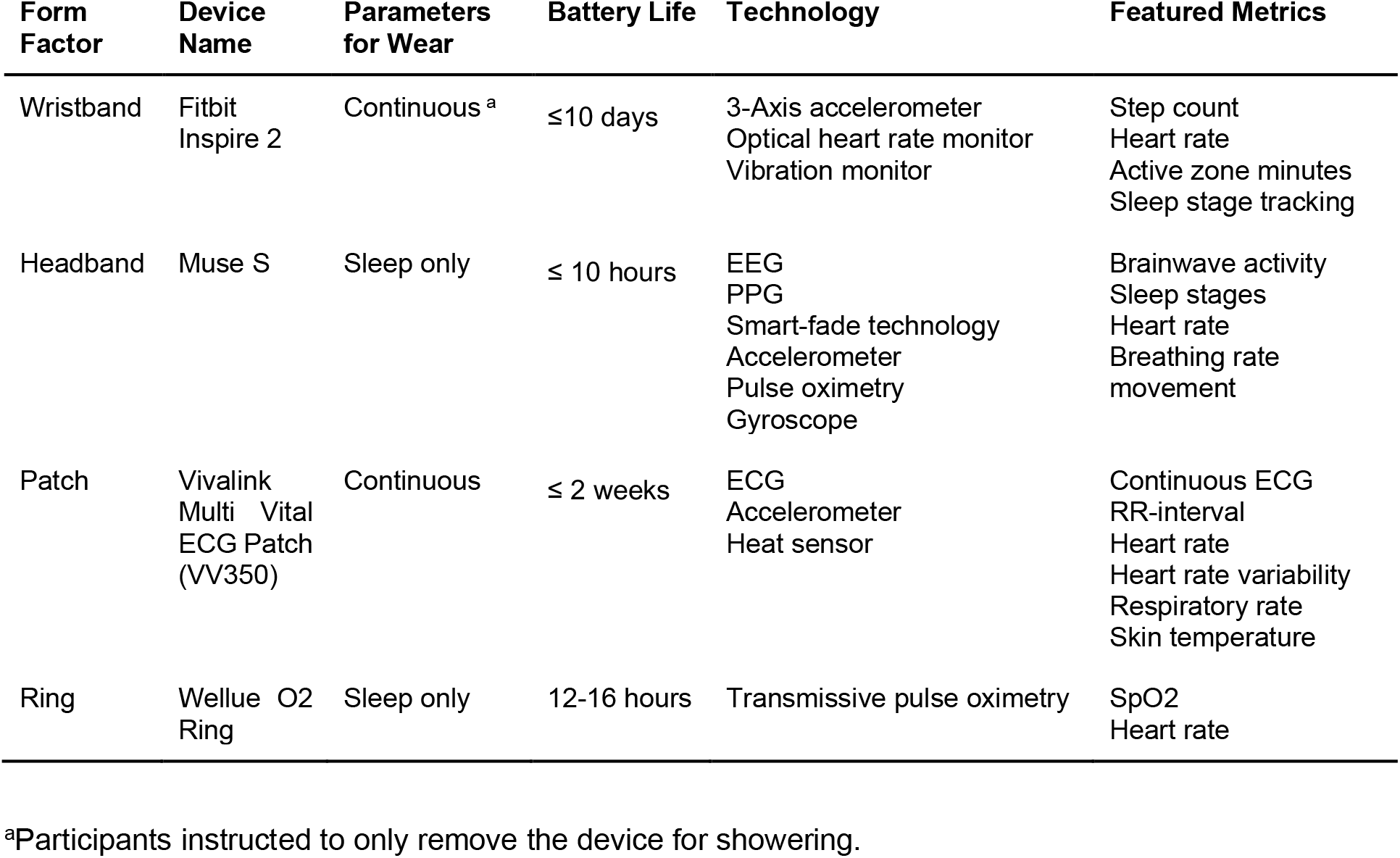
Wearable Device Features.

### Device Protocol Adherence Metrics

To quantify device adherence and data availability, we defined two complementary metrics: completeness and complete expected periods. Completeness was calculated as the percentage of minutes containing valid physiological data relative to the expected duration of monitoring for each device. “Expected duration” differed by device type based on intended wear patterns: 24 hours per day for the wristband and adhesive patch, and 8 hours per day for the headband and ring. For devices intended primarily for sleep monitoring, days with more than 8 hours of valid data were considered 100% complete.

To further characterize adherence in a temporally structured manner, we defined complete expected periods as a discretized measure of data availability. For devices meant to be worn 24h (wrist worn, adhersive chest patch), time was divided into a daytime period (08:00–22:00) and a nighttime period (22:00–08:00). A period was considered complete if the number of minutes with valid data exceeded predefined thresholds corresponding to meaningful coverage of that interval, defined as 600 minutes (10 hours) for the daytime period and 360 minutes (6 hours) for the nighttime period. For devices intended primarily for sleep monitoring (headband and ring), a minimum of 6 hours of valid data within a 24-hour window was considered a complete recording period. Together, these metrics provided both continuous and discretized measures of adherence and data completeness across device types.

### Statistical Analysis

Statistical analyses were conducted to compare adherence and data completeness across device form factors. Because completeness metrics were not assumed to follow a normal distribution, non-parametric methods were used throughout. Group-level differences were first evaluated using the Kruskal–Wallis H test as an omnibus test of distributional differences across device groups. When the omnibus test was significant, pairwise comparisons were performed using two-sided Mann–Whitney U tests to identify specific group differences. To account for multiple comparisons, p-values from pairwise tests were adjusted using the Benjamini–Hochberg procedure to control the false discovery rate. Statistical significance was defined as an adjusted p-value < 0.05.

## Data Availability

Deidentified data can be provided by the corresponding author Aaron D. Besterman upon reasonable request.

## Acknowledgements

This work was supported by a GEMSTONE award from UCSD Institute for Engineering in Medicine (IEM) award number 2000920.

## Author contributions

C.S. performed data analysis, figure generation, and manuscript preparation. A. A. led patient recruitment and training, performed data collection, and aided in manuscript preparation. J.T., K.K., V.G., C.B., and C.R-M., assisted with patient recruitment and training. B.S. advised the data analysis and performed manuscript preparation. A.D.B. oversaw the trial and performed manuscript preparation.

